# Drug Gene-Set Analysis Identifies Established and Emerging Therapeutic Mechanisms

**DOI:** 10.1101/2022.09.06.22279660

**Authors:** Nathaniel Y. Bell, Emil Uffelmann, Eva van Walree, Marijn Schipper, Karl Heilbron, Christiaan de Leeuw, Danielle Posthuma

**Affiliations:** Department of Complex Trait Genetics, Center for Neurogenomics and Cognitive Research, Amsterdam Neuroscience, Vrije Universiteit Amsterdam, Amsterdam, The Netherlands; Department of Psychiatry, Amsterdam UMC, The Netherlands; Department of Child and Adolescent Psychiatry and Pediatric Psychology, Section Complex Trait Genetics, Amsterdam Neuroscience, Vrije Universiteit Medical Center, Amsterdam, The Netherlands; Emma Center for Personalized Medicine, Amsterdam University Medical Center, Amsterdam, the Netherlands; Department of Human Genetics, Amsterdam Reproduction & Development, Amsterdam University Medical Center, Amsterdam, the Netherlands; Stanley Center for Psychiatric Research, Broad Institute of MIT and Harvard, Cambridge, MA, USA; Analytic and Translational Genetics Unit, Department of Medicine, Massachusetts General Hospital, Boston, MA, USA; Department of Psychiatry and Psychotherapy, Charité – Universitätsmedizin Berlin, Berlin, Germany; German Center for Mental Health (DZPG), partner site Berlin/Potsdam, Berlin, Germany; Bayer AG, Research & Development, Pharmaceuticals, Berlin, Germany

**Keywords:** Genome-wide association study, drug repurposing, drug target prioritization, gene-set analysis, psychiatric genetics

## Abstract

Developing novel disease therapies is challenging, especially for central nervous system (CNS) disorders, which have a clinical approval rate of 6.2%. Genetic evidence is associated with higher success rates in drug development, but systematically translating genome-wide association study (GWAS) signals into actionable therapeutic hypotheses remains difficult. Here, we present DRUGSETS, a framework that tests whether drug (*n* = 8,136) and mechanism-of-action (MoA; *n* = 384) gene sets are enriched for GWAS associations. We applied DRUGSETS to five CNS and five non-CNS phenotypes using competitive gene-set analysis with both the standard MAGMA gene-level statistic and Polygenic Priority Scores (PoPS). Both implementations were enriched for approved treatments, and drug gene set significance was associated with further progression in clinical trials. Across all ten phenotypes, we identified 69 MoA and 589 drug gene set-associations that were study-wide significant, finding both established and novel pharmacological targets. Non-CNS traits were enriched for immune, hemostatic and metabolic mechanisms. Psychiatric disorders converged on mechanisms regulating neuronal excitation/inhibition, and Alzheimer’s disease was enriched for immune-related mechanisms and amyloid aggregation. DRUGSETS identifies shared and disease-specific therapeutic biology, including mechanisms not targeted by current treatments. DRUGSETS is available as an open-source tool and is being integrated into FUMA.

## INTRODUCTION

Developing novel pharmaceutical compounds is slow, expensive, and prone to failure: bringing a new medication to market takes an average of 8.3 years and costs $879.3 million, and around 90% of candidates that enter clinical testing never reach patients^1,2^. Attrition is highest where disease biology is least understood, and central nervous system (CNS) conditions are affected most acutely^3^: 13.3% of non-CNS drugs that entered Phase I clinical trials reached market approval, versus just 6.2% for CNS medications^3^. These challenges caused several major pharmaceutical companies to scale back or close their CNS drug discovery programs entirely^4^. Against this backdrop, human genetics provides a promising avenue for improving drug development efforts^5,6^. Therapeutics whose targets are supported by genetic evidence are roughly two times more likely to gain approval, and an estimated 63% of drugs approved between 2013 and 2022 had such support^7–9^.

Genome-wide association studies (GWAS) have identified thousands of genetic loci associated with complex traits and diseases, providing an extensive resource of human genetic evidence for drug development^10,11^. Translating these trait-associated variants into therapeutic hypotheses, however, remains a bottleneck. Locus-to-gene mapping methods nominate effector genes in a single locus but cannot detect mechanisms whose genetic evidence is spread modestly across associated target genes in different loci^12–14^. Mendelian randomization (MR) and transcriptome-wide (TWAS) and proteome-wide association studies can identify candidate target genes but depend on quantitative trait locus (QTL) resources that remain limited in the genes and tissues they cover, and that systematically miss trait-relevant genes^15–18^. Moreover, previous work has shown MR and TWAS do not improve prediction of effector genes^19^. These approaches also evaluate genes one at a time, which makes systematic screening of existing compounds laborious and does not consider that many drugs act on multiple targets.

Drug gene-set analysis, introduced by De Jong et al. (2016), provides a complementary, systematic alternative^20,21^. Each drug is represented by the set of its known protein targets, and a competitive gene-set test asks whether those targets are more strongly associated with the trait than all other genes^20^. Since thousands of drugs can be tested in a single analysis, this enables GWAS results to be used for rapid screening of existing therapeutics. In schizophrenia and bipolar disorder, drug gene-set analysis has recovered antipsychotics and other nervous-system drugs as the most enriched drug classes^21,22^. It has also implicated anticonvulsant targets in bipolar disorder, antidepressant targets in major depressive disorder and GABA-A receptor targets in externalizing behaviours^23–25^.

While these previous studies show the value of drug gene-set analysis, there are two current limitations that leave room for improvement. First, previous implementations have almost exclusively relied on positional gene-level statistics (*e.g.,* MAGMA *z*-scores) to quantify gene importance. This is a powerful method; however, it evaluates each gene in isolation based on variants physically mapped to a locus. Consequently, it overlooks the broader transcriptomic context: whether a gene shares expression patterns across tissues and cell types with other trait-associated genes. While MAGMA *z*-scores capture the association signal at a gene, gene prioritization methods that incorporate this transcriptomic information identify known effector genes more accurately at resolved loci^12^. Second, drugs are tested as individual compounds, whereas GWAS-based gene-set analysis does not give a biological direction of effect. It can establish that a set of genes is associated with a phenotype, but not whether activation of that pathway is risk decreasing or risk increasing^26^. Therefore, compounds acting on the same targets in opposite directions, such as an agonist and an antagonist of the same receptor, will receive near-identical results.

In this study, we address both limitations with DRUGSETS, an open-source Python framework for drug-and mechanism-level gene-set analysis. First, alongside MAGMA gene-level statistics, we also use Polygenic Priority Scores^12^ (PoPS), which learn the expression features that predict gene-level association across the genome, and score genes accordingly. This prioritizes genes by their similarity to trait-associated genes rather than by local signal alone. Second, we assemble mechanism-of-action (MoA) gene sets from ChEMBL^27^ annotations, with action types (*e.g.*, agonist or antagonist) removed, and curate these into broader mechanism groups. This allows the user to test whether a target mechanism is enriched for GWAS signal without assuming a direction of effect, which aligns with the unsigned nature of GWAS-based gene-set analysis. We test whether drug and MoA gene-set associations recover approved treatments greater than expected by chance, and whether higher-ranked drugs progressed further in clinical trials across ten phenotypes. DRUGSETS is available as a standalone software package and is being integrated into the FUMA platform^28^.

## RESULTS

We harmonized drug-target annotations from four public databases: Open Targets^13^, ChEMBL^27^, the Clue Repurposing Hub^29^ and the Drug Gene Interaction Database^30^ (DGIdb). This yielded 8,151 total drug gene sets (*n_genes_* ≥ 2); all drugs were retained regardless of approval status, and DGIdb annotations were filtered at the 50^th^ percentile of the interaction score (see Methods; **Fig. 1a**). Additionally, we defined the direction-agnostic MoA gene sets exclusively using ChEMBL, rather than all four databases, to ensure accuracy of MoA-gene annotations (**Fig. 1b**). Since many ChEMBL MoA annotations consist of only one or a few genes, we manually curated a two-tier hierarchy (Methods). Where possible, related MoAs were grouped into larger parent sets to retain small annotations and increase power, while larger, distinct MoAs were kept as their own parent sets. MoAs with at least two genes within each parent were retained as child sets, representing more specific mechanisms nested within the broader parent groupings (204 parent MoA sets, median set size = 5 genes; 183 child sets, median set size = 2 genes; **Table S1**). After excluding 15 drug and 3 MoA sets that that showed systematic bias regardless of phenotype (Methods), 8,136 drug and 384 MoA gene sets were tested.

**Figure 1.**
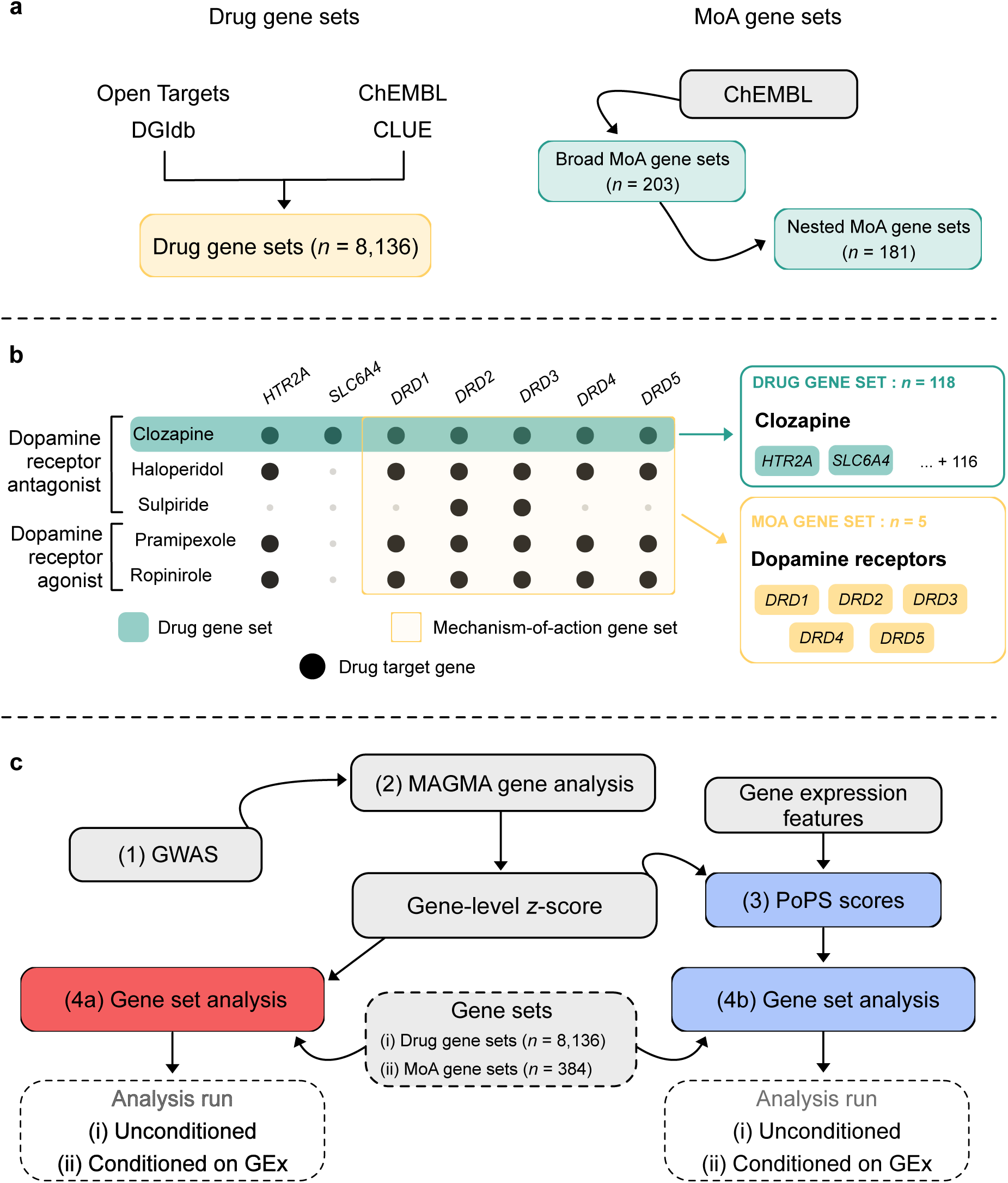
Overview of methods and gene sets. **a,** Drug gene sets (*n* = 8,136) were constructed by harmonizing drug-target annotations from four public databases (Open Targets, ChEMBL, the Clue Repurposing Hub, and DGIdb). Direction-agnostic mechanism-of-action (MoA) gene sets were defined separately from ChEMBL alone, yielding 203 parent MoA sets and 181 child sets. **b,** Example difference between drug and MoA gene sets. **c,** Phenotype-gene set associations were tested using competitive gene-set analysis in MAGMA under two implementations. Gene-level statistics were estimated from GWAS summary statistics with MAGMA. The standard implementation used these *z*-scores directly; the alternative replaced MAGMA *z*-scores with normalized Polygenic Priority Scores (PoPS). Both were run unconditioned and conditioned on gene expression.

To test gene set associations, we used the standard MAGMA competitive gene set analysis with MAGMA *z*-scores. We extended this method to also produce calibrated results when using PoPS scores (see Methods; **Fig. 1c**). For each approach, individual drug gene sets and MoA gene sets (parent and child) were tested for the following phenotypes: asthma^31^, coronary artery disease^32^ (CAD), inflammatory bowel disease^33^ (IBD), LDL cholesterol^34^ (LDL), type 2 diabetes^35^ (T2D), ADHD^36^, Alzheimer’s disease^37^ (AD), bipolar disorder^23^ (BIP), major depressive disorder^24^ (MDD), and schizophrenia^38^ (SCZ; **Table 1**).

**Table 1.** GWAS summary statistics used.

| Phenotype | Cases (n) | Controls (n) | SNP-$h^2$ | Loci | PMID |
| --- | --- | --- | --- | --- | --- |
| **Non-CNS phenotypes** |  |  |  |  |  |
| Asthma | 64,358 | 239,321 | 8% | 167 | 32296059 |
| Coronary artery disease (CAD) | 122,733 | 424,528 | 27.8% | 75 | 29212778 |
| LDL cholesterol (LDL) | 1,320,016 |  | 12%<sup>a</sup> | 403<sup>b</sup> | 34887591 |
| Type 2 diabetes (T2D) | 80,154 | 853,816 | *Not reported* | 193 | 35551307 |
| Inflammatory bowel disease (IBD), meta-analysis<sup>c</sup> | — | — | 11.86% | 84 | 41308126 |
| *IBD* | 21,155 | 753,348 | — | — | — |
| *Crohn's disease* | 9,001 | 752,352 | — | — | — |
| *Ulcerative colitis* | 14,126 | 752,154 | — | — | — |
| **CNS phenotypes** |  |  |  |  |  |
| Attention deficit / hyperactivity disorder (ADHD) | 38,691 | 186,843 | 14% | 27 | 36702997 |
| Alzheimer's disease (AD) | 159,945<sup>d</sup> | 2,409,556<sup>d</sup> | 16% | 114 | 41282776 |
| Bipolar disorder (BIP) | 131,969 | 2,322,416 | 22%<sup>e</sup> | 229 | 39843750 |
| Major depressive disorder (MDD) | 525,197 | 3,362,335 | 8.4% | 570 | 39814019 |
| Schizophrenia (SCZ) | 53,386 | 77,258 | 24% | 263 | 35396580 |

<sup>a</sup>Reported estimate derived from genome-wide significant SNPs only.
<sup>b</sup>Number of genome-wide significant lead SNPs for EUR ancestry; ancestry-specific locus counts were not reported.
<sup>c</sup>The IBD GWAS is a meta-analysis across three case definitions.
<sup>d</sup>Case and control counts include proxy cases and controls based on parental diagnosis.
<sup>e</sup>Percentage reflects SNP- $h^2$ computed from clinically defined case definitions.

### Benchmarking drug and MoA gene set analyses

To jointly compare the standard MAGMA *z*-score and PoPS-modified drug gene set implementations, we assessed their recovery of known therapeutic target biology across the ten phenotypes for both individual drug and MoA gene sets. For each phenotype, a permutation test compared the mean rank by *P* value of drugs and MoAs approved for that indication against 10,000 permutations (see Methods).

At the drug level, both implementations recovered approved compounds. PoPS-based results reached FDR significance for nine of ten phenotypes, while the MAGMA *z* results were FDR significant for eight of ten (**Fig. 2a**). We observed weaker per-phenotype recovery for the MoA gene sets (2/10 FDR-significant, with an additional three reaching uncorrected significance for PoPS; 2/10 FDR-significant for MAGMA *z*, with an additional two reaching uncorrected significance; **Fig. 2b**; Supplementary Table 3). Fisher exact tests confirmed global enrichment across all phenotypes for both individual drug and MoA levels and for both implementations (drugs: *P_MAGMA_* = 2.3 × 10^-9^, *P_PoPS_* = 3.66 × 10^-4^; MoA: *P_MAGMA_* = 5.02 × 10^-4^, *P_PoPS_* = 7.86 × 10^-5^).

**Figure 2.**
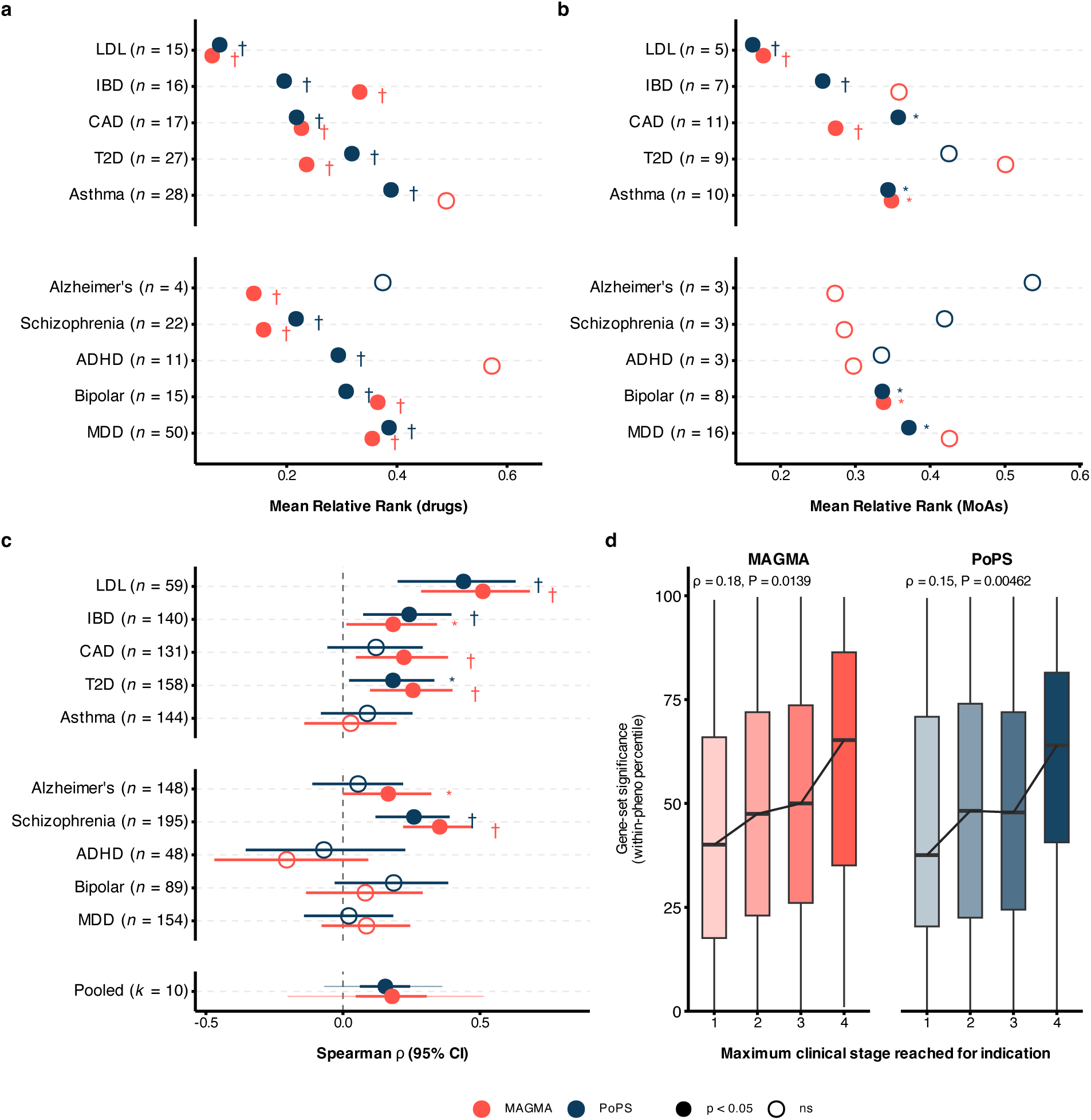
Standard MAGMA and PoPS implementations recover known drugs and targets. **a, b,** Dot plots showing the observed mean relative rank of approved drugs **(a)** and MoA **(b)** gene sets for all ten phenotypes. Significance was determined by comparing observed mean ranks of approved sets to 10,000 means of randomly drawn sets of equal size (see Methods). Lower rank indicates more accurate prioritization of established therapeutic biology. Numbers next to phenotype names indicate the number of approved gene sets for that trait. **c,** Forest plot of Spearman rho estimates between effective gene set significance -log10(*P*) and attained clinical phase per phenotype, with bars representing 95% confidence intervals. Pooled estimate reflects random-effects meta-analysed Spearman rho across all phenotypes, with thick bars representing 95% confidence intervals and thin bars representing 95% prediction intervals. **d,** Box plots of within phenotype drug gene set significance across clinical trial phases. * = *P* < 0.05; † = FDR significance (*PFDR* < 0.05). For all panels, vermilion = MAGMA; indigo = PoPS.

We next examined if drug sets ranked higher by *P* value were more likely to have progressed further in clinical trials. Spearman’s rho was positive in nine of ten phenotypes, with five phenotypes having at least one of MAGMA *z* or PoPS-based results reaching FDR-corrected significance (**Fig. 2c**). A random-effects meta-analysis supported a positive pooled effect across phenotypes (pooled rho = 0.18, *P* = 0.013 for MAGMA *z*; pooled rho = 0.15, *P* = 0.004 for PoPS; **Fig. 2d**; **Table 2**).

**Table 2.** Meta-analysis of association between gene-set significance and attained clinical phase.

| Analysis | K | Rho | 95% CI | P | 95% PI | tau<sup>2</sup> | I<sup>2</sup> | Q | P<sub>q</sub> |
| --- | --- | --- | --- | --- | --- | --- | --- | --- | --- |
| MAGMA z | 10 | 0.179 | 0.047-0.306 | 0.013 | -0.202 – 0.513 | 0.026 | 74.6 | 26.82 | 0.001 |
| PoPS | 10 | 0.154 | 0.061-0.245 | 0.004 | -0.07 – 0.363 | 0.008 | 48.1 | 15.54 | 0.077 |

Random-effects meta-analysis of per-phenotype Spearman's rho between gene-set significance and maximum attained clinical phase. Analysis, gene statistics used as input to MAGMA competitive gene-set analysis; K, number of phenotypes; 95% PI, prediction interval for the true effect in a new phenotype; $\tau^2$ , between-phenotype variance on the Fisher z scale; I<sup>2</sup>, percentage of total variation attributable to between-phenotype heterogeneity; P<sub>q</sub>, P value from Cochran's Q test of homogeneity. P values are two-sided.

### Mechanism gene sets recover known pharmacology and nominate novel targets

In total, 384 MoA gene sets were tested across the ten phenotypes, yielding 69 significant gene set associations after study-wide FDR correction (Supplementary Table 4). We use the five non-CNS traits as a positive control for the MoA gene set analysis. Recovering the pharmacology of these traits serves as a validation, since the therapies for these disorders act on well characterized target mechanisms. Across all non-CNS traits, the MoA gene set analysis recovered at least one pharmacological target of an approved medication at study-wide FDR significance for their respective indications.

For asthma, we identified two study-wide FDR significant MoA sets targeted by approved therapeutics: interleukin receptor and interleukin cytokine signalling (**Fig. 3a**). Alongside these approved targets, we observed strong associations between asthma and the novel target mechanism sets of T-cell signalling, the TNF-receptor and ligand superfamilies, and the NF-*κ*B pathway (**Fig. 3a**). For IBD, one MoA gene set targeted by approved treatments reached study-wide significance: integrin α4β7 (target of vedolizumab; **Fig. 3b**). CAD and LDL cholesterol converged on hemostatic mechanisms, with the coagulation pathway reaching study-wide significance for both phenotypes (**Fig. 3c**). Many other MoA sets related to vascular biology displayed significant associations with CAD, including the angiopoietin-Tie pathway, fibrinolytic system, endothelin receptor family, and VEGFR/PDGFR/KIT kinase sets (**Fig. 3c**). For LDL cholesterol, the mevalonate/HMG-CoA (target of statins), microsomal triglyceride transfer protein (MTP), and lipid nuclear receptors – all targeted by approved treatments for LDL – were study-wide significant (**Fig. 3d**). In T2D, we observed significant associations between the approved target ATP-sensitive potassium (K^+^_ATP_) channels and the highly related specific potassium channel configuration SUR1–Kir6.2 (**Fig. 3d**). Promising unapproved target mechanisms related to cell-cycle machinery regulating pancreatic β-cell proliferation, including the CDK6–cyclin D1 and CDK4–cyclin D1 complexes, along with Notch and phosphodiesterase-3 signalling sets were also associated with T2D. Overall, the recovery of validated pharmacology across the non-CNS traits demonstrates that the MoA gene set analysis can identify real, druggable biology using GWAS signal.

**Fig. 3.**
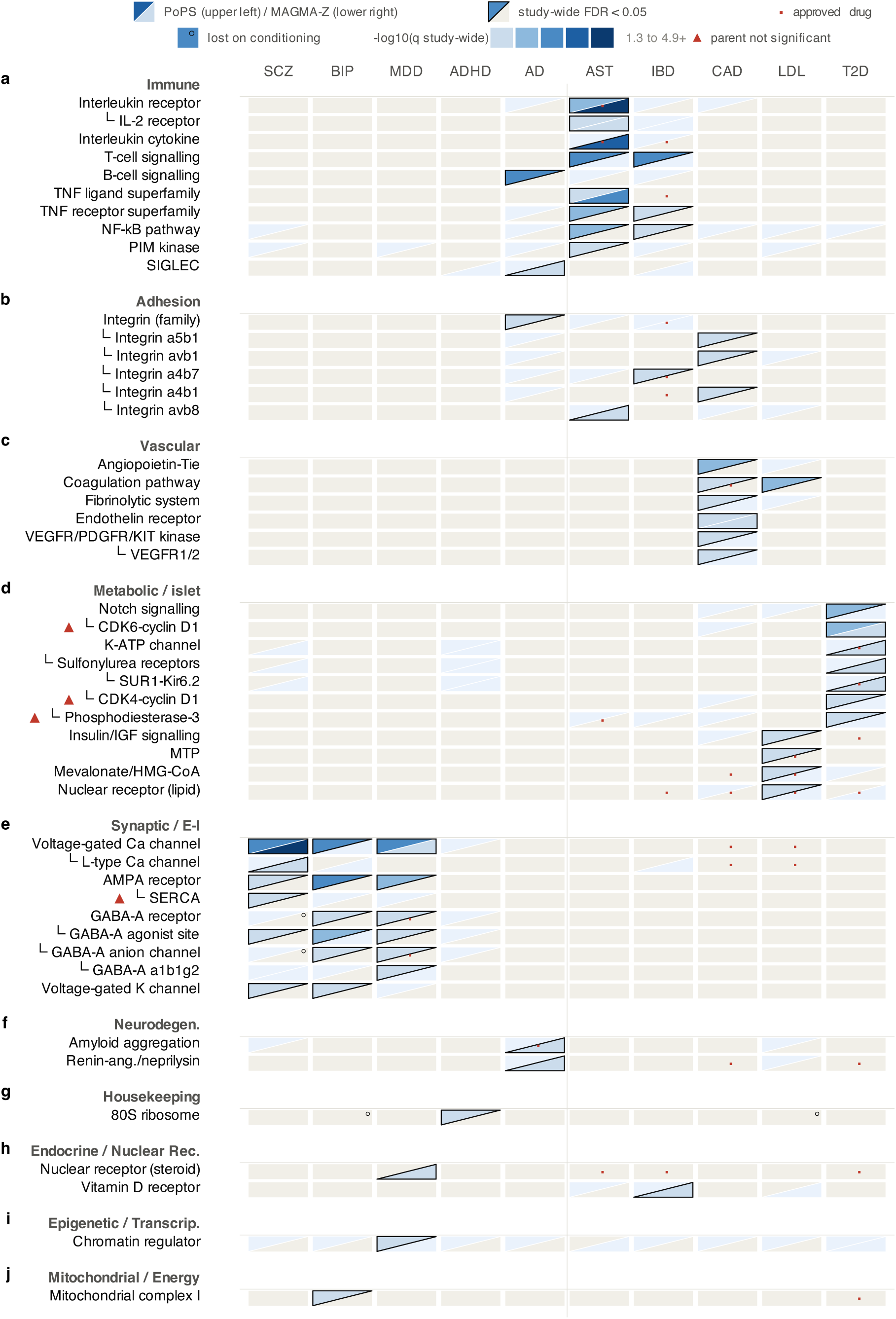
Mechanism of action gene set analysis results. Heatmap of mechanism-of-action (MoA) gene set analysis results across all phenotypes for the MAGMA *z* and PoPS-based implementations. Displayed gene sets reached study-wide FDR significance for at least one phenotype. All results displayed are from competitive gene set analysis performed while conditioning on gene expression (see Methods). Cells are colored by -log10(*P*) of their MoA gene set, with upper left color representing PoPS-based results and bottom right representing MAGMA *z*-based results. Solid outline signifies FDR study-wide significance. Sets are grouped together according to biological function. └─ indicates a direct child gene set of the parent set above it; red triangle = an orphan child set (*i.e.*, parent set was not significant). Circle in upper right = set was significant in the analysis without conditioning on gene expression; red dot = approved target for phenotype (in ChEMBL data).

In contrast to the immune, coagulation, and metabolic mechanisms recovered in the non-CNS traits, we observe shared and distinct target biology centered on the regulation of neuronal excitation/inhibition (E/I) balance for MDD, BIP, and SCZ. Voltage-gated calcium channels, the GABA-A receptor agonist site, and AMPA-type glutamate receptor sets all reached study-wide significance for the three psychiatric traits (**Fig. 3e**). Additionally, the parent GABA-A receptor and the child GABA-A anion channel sets were both study-wide significant for BIP and MDD, and the voltage-gated K^+^ channel set was significant for SCZ and BIP (**Fig. 3e**). We also observed some phenotype-specific associations with the psychiatric phenotypes: the sarcoplasmic/endoplasmic reticulum calcium-ATPase (SERCA) was study-wide significant for SCZ, as was the specific GABA-A α1β1γ2 subunit configuration set for MDD (**Fig. 3e**). Taken together, these MoA sets span the core machinery of synaptic signalling: glutamatergic excitation (AMPA), GABAergic inhibition (GABA-A), and the voltage-gated calcium and potassium channels that regulate neurotransmitter release and membrane repolarization. These findings implicate E/I homeostasis as a druggable domain and mark it as a candidate for therapies that act on shared psychiatric biology.

The remaining CNS traits, AD and ADHD, departed from this shared E/I axis. AD displayed strong associations with immune-related MoA sets: B-cell signalling, the integrin family (including the specific αIIbβ3 heterodimer set), and SIGLEC receptors, implicating innate and adaptive immune signalling. This aligns with known involvement of microglial and immune system activity in AD^37^. Furthermore, AD also displayed significant associations with the amyloid aggregation (target mechanism of lecanemab) and the renin-angiotensin/neprilysin MoA sets (**Fig. 3f**). ADHD, by contrast, yielded less interpretable signals: its study-wide associations were confined to the 80S ribosome set (**Fig. 3g**). While ribosomal proteins have been implicated for ADHD previously, their tractability as pharmacological targets is unclear^39,40^. More promisingly, the dopamine D2/D3 receptor set was the second-ranked MoA for ADHD (uncorrected *P_PoPS_* = 0.002) and was accompanied by dopamine D2-like receptor (uncorrected *P_PoPS_* = 0.008) and dopamine D1/D2 receptor (uncorrected *P_PoPS_* = 0.009) sets in the top five strongest associations with ADHD. Dopamine signalling is a key target for first-line stimulants such as methylphenidate, which block the dopamine transporter to increase dopamine bioavailability.

Considered together, these results show that MoA gene set analysis can recover established pharmacology and nominate biologically coherent novel mechanisms across ten distinct phenotypes. The study-wide significant gene sets for the non-CNS results are driven by immune, coagulation, and metabolic signalling, whereas the CNS traits are split between E/I biology for MDD, BIP, and SCZ, and immune and amyloid aggregation for AD. Where the CNS and non-CNS results converge, they do so along expected mechanistic lines: AD, the included CNS disease with the clearest immune involvement, shares immune MoA gene sets with the immune-mediated non-CNS traits, while the psychiatric E/I axis shows no association with non-CNS phenotypes. That both this overlap and the broader separation between domains track known biology suggests these associations reflect genuine signal rather than generic druggability.

### Compound and class-level results

We next tested 8,136 individual drug gene sets using both the MAGMA *z* and PoPS-based implementations (see Methods). In total, 589 associations reached FDR study-wide significance and were unevenly distributed across phenotypes, ranging from 86 for IBD to five for ADHD. To prioritize the strongest repurposing candidates, we restricted the following results to compounds with existing regulatory approval for at least one clinical indication (clinical phase 4), with full results available in Supplementary Table 5.

The drug gene-set analysis of non-CNS phenotypes successfully prioritized established and highly relevant treatments. We found at least one approved treatment in the top five ranked compounds for asthma (beclometasone), CAD (alirocumab), LDL cholesterol (lomitapide; rosuvastatin; mipomersen), and T2D (repaglinide; exenatide; glipizide; sitagliptin; **Fig. 4a**). Beclometasone was significant for IBD, which is not flagged as approved in the ChEMBL data but is an approved IBD treatment in Europe^41^ (**Fig. 4a**). We also identified several candidate drug repurposing compounds that were not approved for their respective phenotypes. The anti-CD25 antibody daclizumab, which demonstrated moderate improvement in a phase 2 clinical trial in asthma patients, was strongly associated with both asthma and IBD^42^ (**Fig. 4a**). For LDL cholesterol and CAD, both traits recovered drugs affecting hemostasis, reflecting the coagulation and fibrinolytic MoA pathways identified in **Fig. 3c/d**: tranexamic acid (CAD), antithrombin alfa (LDL), heparin calcium (CAD and LDL), and the thrombopoietin receptor agonists avatrombopag and lusutrombopag (LDL; **Fig. 4a**). The VEGFR inhibitor tivozanib was also significant for CAD, consistent with the VEGFR/PDGFR/KIT kinase MoA finding in **Fig. 3c**. In T2D, the dual PPAR α/δ agonist elafibranor ranked second among all sets tested. Originally developed for T2D and hyperlipemia, elafibranor is now used for cholangitis and is being tested in clinical trials for cirrhosis and liver disease^43^ (**Fig. 4a**).

**Fig. 4.**
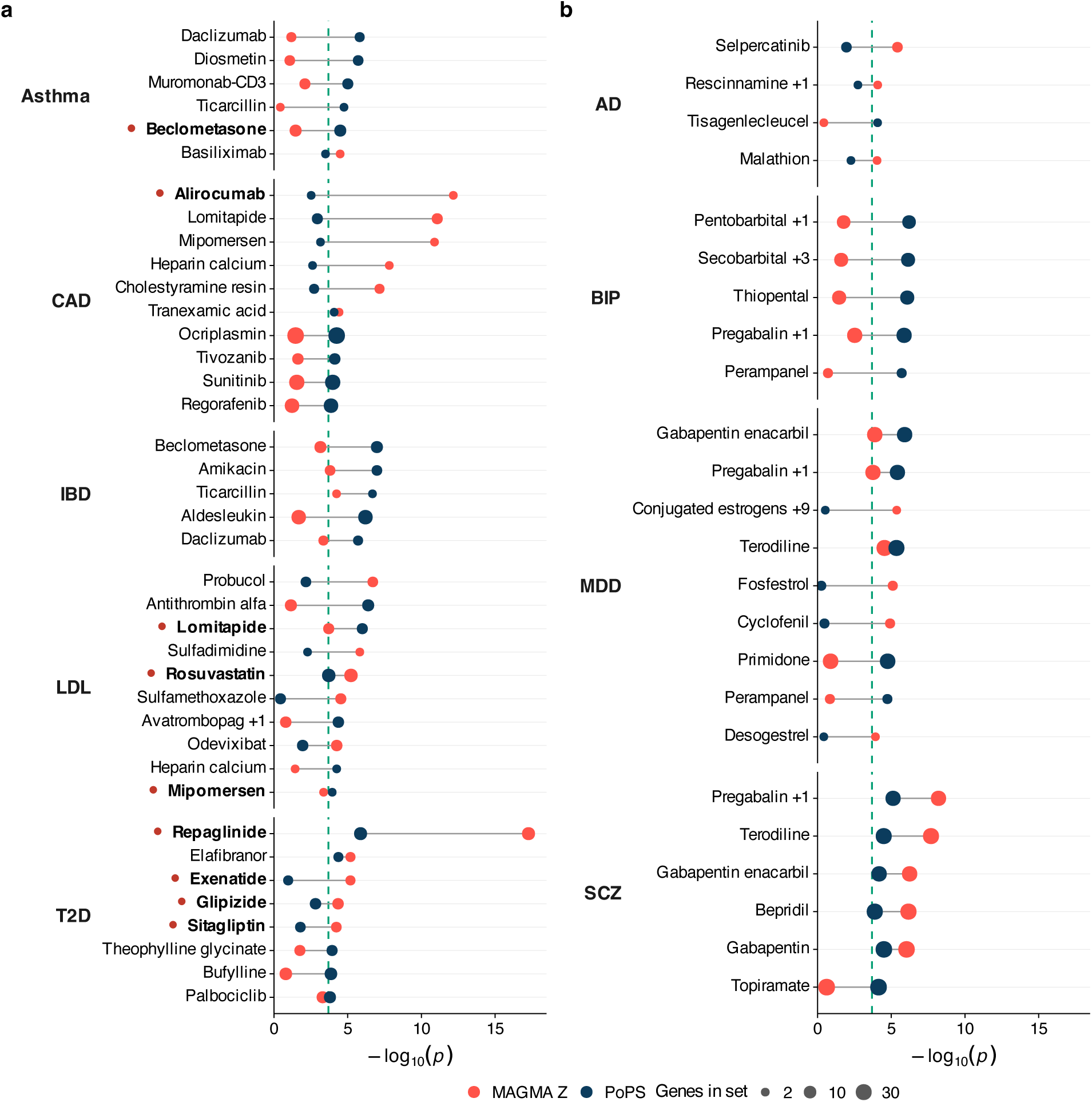
Drug gene set analysis results for approved compounds. Dumbbell plots displaying individual drug gene-set analysis results across non-CNS **(a)** and CNS **(b)** phenotypes for the MAGMA *z* and PoPS-based implementations. Displayed gene sets represent up to the top five significant compounds per phenotype for each method that reached FDR-corrected study-wide significance, restricted to compounds that have reached clinical approval for any disease. The points represent the competitive gene set analysis -log10(*P*) of the drug gene set, colored by method. Point size corresponds to the number of genes in the set. Green vertical dashed line represents study-wide FDR significance threshold. Drug names are bolded and marked with a red circle if the compound is approved for its phenotype. Where multiple drugs share an identical gene set, a single drug is named alongside a count of the additional gene sharing compounds (e.g., +1).

In contrast to the non-CNS phenotypes, the CNS phenotypes results contained no drugs approved to treat the diseases they were associated with (**Fig. 4b**). The leading AD compounds differed between the two analysis types, but neither analysis returned amyloid-acting drugs. The MAGMA *z* analysis identified the RET inhibitor selpercatinib (*P_FDR_* = 4.2 × 10⁻³), the rauwolfia alkaloids deserpidine and rescinnamine (*P_FDR_* = 0.035), and the acetylcholinesterase inhibitor malathion (*P_FDR_* = 0.037). The malathion result is notable, as acetylcholinesterase inhibition is the mechanism of donepezil and rivastigmine, which are both currently approved to treat AD. The PoPS framework identified only the CD19-directed CAR-T therapy tisagenlecleucel (*P_FDR_* = 0.035). The anti-amyloid antibody lecanemab ranked next in both analyses but did not reach study-wide significance (MAGMA *z P_FDR_* = 0.053; PoPS *P_FDR_* = 0.086). These compound-level results correspond to our MoA-level, and this mechanistic convergence between compound-and MoA-level analyses was also evident in psychiatric traits. The top-ranked compounds for BIP were GABA-A-targeting barbiturates (pentobarbital, secobarbital, thiopental), while MDD and SCZ were primarily associated with α2δ ligands acting on voltage-gated calcium channels (*e.g.*, gabapentin, pregabalin; **Fig. 4b**). Lastly, no ADHD-approved drugs reached study-wide significance, but its strongest hits (imipramine, PoPS *P_FDR_* = 0.076; nomifensine, PoPS *P_FDR_* = 0.091) target the same noradrenaline and dopamine transporters as first-line ADHD treatments, consistent with the dopaminergic MoA sets.

To test for compound-level enrichment within therapeutic classes, we regressed the drug gene-set test statistics on membership of each Anatomical Therapeutic Chemical (ATC) level 3 class, with and without modelling the correlation between drug gene sets (Methods; Supplementary Note 1). The principal treatment class was significant in all four analyses for LDL cholesterol (lipid-modifying agents, C10A), T2D (blood glucose-lowering drugs, A10B) and IBD (immunosuppressants, L04A; Supplementary Fig. 5; Supplementary Table 6). For SCZ and BIP, antipsychotics (N05A), and for BIP antiepileptics (N03A), were significant only when target sharing was not modelled, mirroring the weaker recovery of approved compounds for CNS traits. Among the CNS traits, only one class was significant in all four analyses: hypnotics and sedatives (N05C) for BIP, in line with the barbiturate and GABA-A receptor results above.

### Concordance between implementations and gene set families

Given the two gene-level statistics used in the MAGMA *z*-score and PoPS-based implementations, we examined the relationship between these two sets of results. For each phenotype we correlated the competitive test statistics (β/SE) from the two approaches, separately for MoA and drug gene sets (**Fig. 5a, b**). We observed moderate agreement that was consistent in direction across all ten phenotypes, with a median Pearson *r* of 0.457 for MoA sets (range 0.40 to 0.65) and 0.435 for drug sets (range 0.35 to 0.60). This similar but non-identical relationship is to be expected: MAGMA *z*-scores are used in the PoPS model so the two are correlated by design and should not be treated as independent replications of each other. These findings also underscore that the MAGMA *z* and PoPS-based approaches are complementary.

**Fig. 5.**
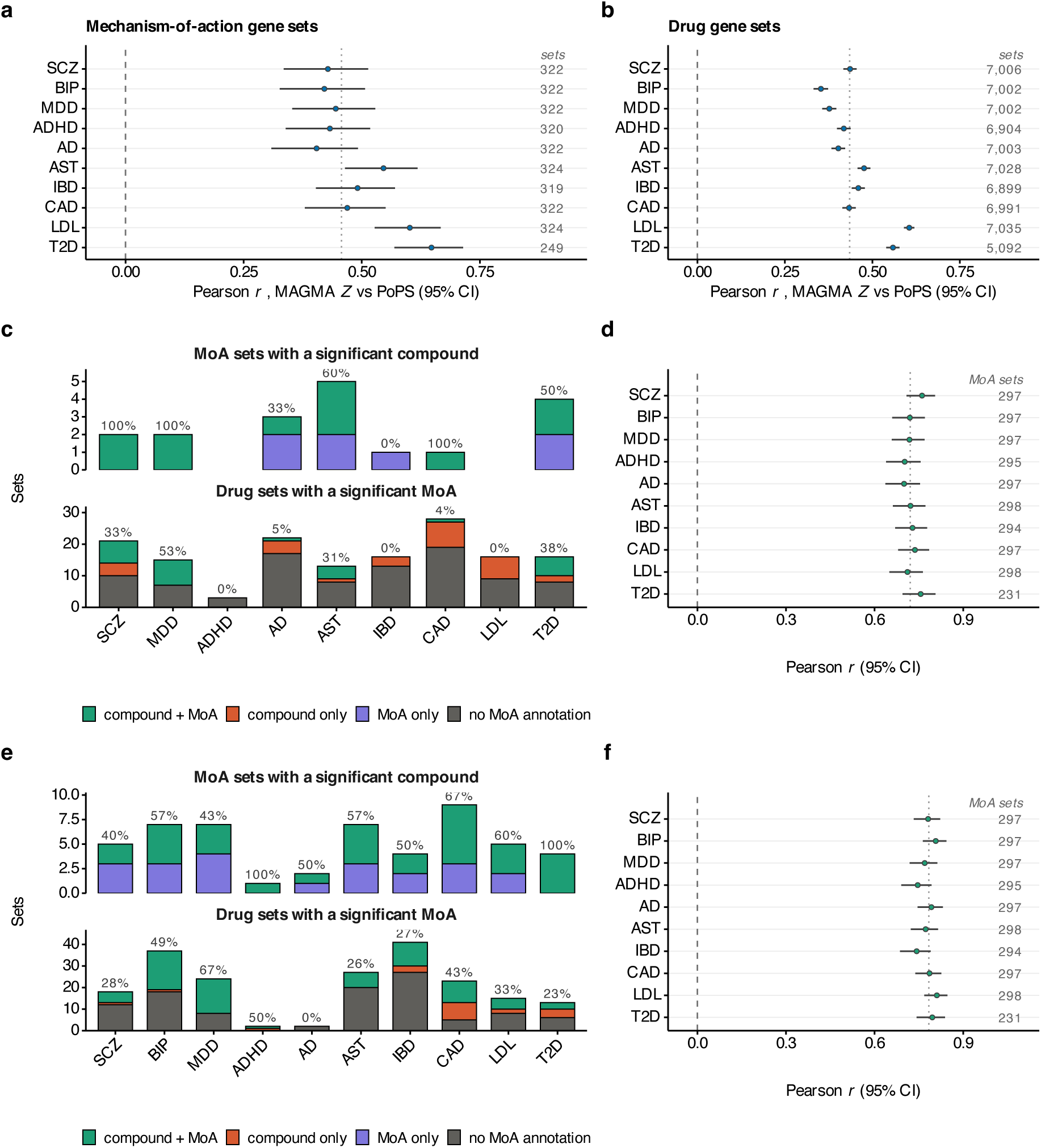
Concordance between implementations and gene set families. **a, b** Within-phenotype correlations between the gene set test statistics (β/SE) for MAGMA *z* and PoPS-based implementations, for MoA sets **(a)** and drug sets **(b)**. Points are correlation estimates, horizontal bars 95% confidence intervals; the dashed line marks zero and the dotted line the median across phenotypes. Sample sizes are given on the right. **c-f** Convergence between drug-level and mechanism-level results for the MAGMA *z* **(c, d)** and the PoPS-based **(e, f)** implementations. **c, e** Significant MoA (top) and drug (bottom) gene sets per phenotype, split by convergence: green, matched MoA and drug sets both significant; orange, significant drug set with non-significant annotated MoA term; purple, significant MoA set with insignificant annotated drug; grey, drug sets with no annotated MoA. Percentages give the proportion that are convergent; empty bars indicate no significant sets for that phenotype. **d, f** Within-phenotype Pearson correlations between the MoA test statistic and the mean test statistic of the drug sets annotated to that mechanism, plotted as in **a** and **b**.

Similarly, given that mechanism-of-action and drug gene-set analyses test related but not identical hypotheses, we asked whether the two approaches recover the same underlying genetic signal. Each drug gene set that had a MoA annotation was paired with that mechanism – with the child mechanism selected if a drug had parent and child MoA annotations – and the significance and competitive test statistics were compared across the ten traits (**Fig. 5c-f**). This resulted in 18,285 compound-MoA pairs across all phenotypes, ranging from 1,368 (T2D) to 1,888 (LDL and asthma) per trait. We observed partial convergence between significant drug sets and MoA sets in both directions. Among the MoA sets that reached significance for a given trait, the proportion for which at least one corresponding drug set was also significant ranged of 0% to 100% for MAGMA *z* results (median = 60%) and from 40% to 100% for PoPS-based results (median = 57%). Conversely, among significant drug sets, an average of 61.5% of MAGMA *z* results and 51% of PoPS-based results across phenotypes had no corresponding MoA annotations (**Fig. 5c, e**). Lastly, we observe highly consistent positive relationships between the mechanism-level gene set test statistics and the mean test statistic of its annotated compounds for every phenotype for both MAGMA *z* (median Pearson *r* = 0.72) and PoPS implementations (median Pearson *r* = 0.784; **Fig. 5d, f**). Thus, while we observe meaningful concordance between drug and MoA gene set associations, the overlap is incomplete. This partial convergence underscores that evaluating both individual compound and MoA target sets provides complementary insights for GWAS-based drug or target prioritization.

## DISCUSSION

In this study, we developed and applied a GWAS-based framework for prioritizing therapeutic compounds and pharmacological mechanisms across ten phenotypes using 8,136 drug and 384 MoA gene sets. Our results were enriched for approved targets and treatments, and stronger drug set significance was associated with further progression in clinical trials. Beyond recovering approved biology, both implementations nominated coherent unapproved mechanisms that align with known disease etiology: immune, hemostatic and metabolic domains for non-CNS traits, and predominantly E/I-related pathways across psychiatric disorders and amyloid and immune processes for AD. This work demonstrates that GWAS signals can be systematically used to identify established and novel pharmacological mechanisms, and we make this framework available as an open-source software tool, DRUGSETS.

Across MDD, BIP, and SCZ the prioritized mechanisms center on the regulation of neuronal excitation/inhibition (E/I) balance. Voltage-gated calcium channels, AMPA-type glutamate receptors, and GABA-A receptors were significant for all three disorders. These mechanisms differ from the current approved treatments for these phenotypes, which act primarily on monoamine systems, such as serotonin and noradrenaline reuptake for MDD, dopamine D2 antagonism for SCZ or, in the case of lithium for BIP, on mechanisms that remain largely uncharacterized^44–47^. Our findings are nevertheless consistent with a substantial body of evidence in a range of brain-related phenotypes. Previous GWASs have repeatedly identified genes in this domain, including calcium channels, AMPA receptors, and GABA receptors^23,24,38^. Additionally, genetic findings align with evidence from research on GABAergic interneuron pathology, EEG beta and gamma-band measures, and animal models of altered E/I ratio, altogether forming a strong hypothesis for the relationship between E/I imbalance and psychiatric phenotypes^48–51^. Our analyses show that this signal concentrates in protein families that existing compounds already target. This is of practical interest, as several therapies targeting these mechanisms have recently entered clinical use. Esketamine is approved for treatment-resistant depression, and racemic ketamine is used off-label for the same indication; their antidepressant effects are thought to depend on downstream AMPA receptor activation^52–55^. Similarly, the GABA-A positive allosteric modulators brexanolone and zuranolone were recently approved for postpartum depression^56–58^. Preclinically, SEAD1, a synthetic version of the voltage-gated Ca^2+^ channel subunit α2δ-1, reversed synaptic and behavioral deficits in a mouse model of SCZ^59^.

A similar pattern was evident for the non-CNS phenotypes. The asthma and IBD results converged on immune mechanisms that are supported by clinical evidence including the TNF superfamily and IL-2/CD25 signalling. TNF inhibitors are approved for IBD, and the anti-CD25 antibody daclizumab (approved for multiple sclerosis) showed efficacy in a phase 2 asthma trial^42^. Moreover, low-dose IL-2 therapy is now under investigation for IBD and has undergone phase 2 clinical trials for patients with allergies and was well tolerated by those with asthma^60,61^. Likewise, our results for CAD and LDL strongly prioritized known treatments and mechanisms, such as the alirocumab drug set (an LDL-lowering antibody therapy) and the coagulation MoA set for CAD, and the rosuvastatin drug set and the HMG-CoA mechanism set for LDL. Our results also identified mechanisms for both that do not yet have approved treatments, including the angiopoietin-Tie signalling, fibrinolytic system, and endothelin receptor MoA sets for CAD, and the insulin/IGF signaling set for LDL. We also see a similar pattern for T2D, with multiple approved compounds (*e.g.*, repaglinide, exenatide, glipizide, *etc*.) and the approved K^+^_ATP_ channel MoA set all being prioritized. All these results taken together provide a body of evidence that the drug gene-set analysis framework applied here can identify pharmacological targets and compounds that are known to work alongside novel targets worth exploring in follow-up research.

Benchmarking drug repurposing methods against approved drugs carries some important interpretational challenges. A method that recovers only approved targets and drugs provides little gain for identifying novel therapeutic approaches. Conversely, methods that identify no approved therapies cannot be easily benchmarked with publicly available data. Our results seem to have struck a balance between these two possibilities, and this balance hinges along the CNS and non-CNS phenotype domain axis. We observed three findings that illustrate this: the enrichment of approved MoA gene sets was limited for CNS phenotypes, the positive relationship between drug set significance and clinical trial progression seemed primarily driven by non-CNS phenotypes (AD and SCZ being the exceptions), and the top-ranked compounds for CNS phenotypes were absent of approved therapies, unlike their non-CNS counterparts. One explanation is that the benchmark itself is weaker for CNS traits. The complex biology underlying CNS phenotypes – paired with additional drug development challenges like the blood brain barrier – has resulted in fewer approved therapies that target the causal biology compared with non-CNS disorders. The rank-based permutation tests point toward this first explanation: between the MAGMA *z*-score and PoPS-based approaches, approved compounds ranked higher than chance for all five CNS phenotypes. Therefore, the genetic signal is not absent for CNS drugs (as indicated by the observed enrichment), but it is currently not strong enough to place them among the top hits. This is consistent with the CNS results converging on E/I and immune mechanisms, which have stronger common-variant GWAS signal than the targets of existing treatments, such as monoamine biology. However, we cannot fully separate these explanations with the current data.

Our results support the beneficial use of GWAS results for drug repurposing and target identification efforts. Nevertheless, the drug and MoA gene set analyses also leave some questions unanswered. Most broadly, these analyses identify mechanisms enriched for genetic associations. However, effective drugs can act downstream of causal biology, targeting symptoms rather than underlying disease mechanisms, and such therapeutics are unlikely to be discovered by drug- or MoA gene-set analysis (*e.g.*, β_2_-adgrenergics for asthma). Second, competitive gene-set analysis has no direction of effect, so drugs identified using this method should be interpreted with caution, as they have the potential to both exacerbate and alleviate disease progression and symptoms. The mechanism-based analysis partially bypasses this by testing targets without assuming a direction of effect. Third, as discussed above, this method identifies drugs and mechanisms based on their genes and does not consider other pharmacologically relevant details. For example, a topically applied dermatological treatment may be prioritized for a phenotype that is typically treated systemically with oral medication. Such a result is not spurious: the compound is prioritized because its targets are strongly associated with the phenotype under investigation. It does, however, illustrate that the gene set analyses executed above are agnostic to formulation, route of administration and tissue. Finally, many drug and MoA gene sets are small: the median parent MoA set contains five genes, and the median child set contains two. Because gene set analysis requires a minimum of two genes, highly specific drugs and mechanisms that act on a single target are excluded from DRUGSETS entirely.

Taken together, the results we present here establish drug and MoA gene set analyses as practical methods for using human genetic evidence for pharmacological prioritization. This approach nominates target biology and drug candidates, with questions of direction of effect, modality, and clinical viability requiring dedicated follow-up. The value of drug and MoA gene set analyses therefore lies at the start of a repurposing pipeline, or as one of several methods used for triangulating novel compounds or targets for a phenotype. To support its use, we provide the full pipeline as an open-source Python command-line tool, DRUGSETS, which is being integrated into the FUMA platform for easy application to GWAS summary statistics. We show that GWAS signal can be used to recover known treatments while simultaneously identifying biologically coherent therapeutic targets beyond existing established treatments.

## MATERIALS & METHODS

### GWAS summary statistics

We obtained publicly available GWAS summary statistics for ten phenotypes, comprising five CNS and five non-CNS phenotypes. To match the LD reference used by MAGMA, We used European-ancestry summary statistics for every phenotype or restricted to the European-ancestry subset where a broader multi-ancestry meta-analysis was available. For all summary statistics, the major histocompatibility complex was removed based on the Genome Reference Consortium definition. For a full overview of summary statistics used, see Table 1.

### Construction of individual drug gene sets

We created a harmonized drug–target repository from four public resources: ChEMBL^27^ (v36); Open Targets^13^ (v26.03); Clue^29^ (2020 release); and the Drug–Gene Interaction Database^30^ (DGIdb; accessed 21 May 2026). We used ChEMBL identifiers to map compounds between databases, and linked Clue compounds to ChEMBL identifiers via their PubChem CIDs using UniChem cross-references. If a compound had no ChEMBL identifier across all datasets, its drug name was used instead. Child molecules (*e.g.,* different salt forms) were merged to their parent compound using ChEMBL molecule-hierarchy annotations. Gene-level targets were drawn from all four sources: DGIdb (drug-gene interaction records), Open Targets (target genes from its known-drug dataset), Clue (Repurposing Hub target annotations), and ChEMBL. For ChEMBL, drug-target assignments were taken from the manually curated “drug_mechanism” table. We note that Open Targets extracts its drug-target annotations from ChEMBL, so there was substantial overlap between these two sources; we use both for thoroughness. Mechanism-of-action labels, ATC classes, and clinical indications (with their per-indication approval phase) were supplied by ChEMBL wherever possible. Gene sets were defined as the union of all target genes mapped to a compound across the four sources. Since DGIdb drug-target interactions are accompanied by confidence scores, we assessed four approaches for adding DGIdb genes to gene sets: (i) including all interactions; filtering out genes below the (ii) median (0.136) or (iii) 75^th^ percentile (0.678) interaction score; and (iv) excluding DGIdb entirely. In parallel, we constructed drug gene-set collections at successive maximum approval phase thresholds, ranging from all compounds to approved compounds only. For all analyses here, we use the median DGIdb cut-off (to ensure confidence of drug-gene annotations) and retain all compounds regardless of approval status. However, all combinations of drug gene sets are available within the DRUGSETS framework.

### Construction of mechanism-of-action gene sets

Direction-agnostic mechanism-of-action (MoA) gene sets were constructed exclusively from ChEMBL (v36). Drug-target mechanisms were extracted from ChEMBL’s “drug_mechanism” table, and UniProt human protein identifiers were translated to Ensembl gene IDs using org.Hs.eg.db. The action type (*e.g.*, agonist, antagonist) was removed from the MoA label so that pharmacologically opposed activities collapsed onto shared molecular targets (*e.g.*, GABA-A receptor agonists and antagonists both map to GABA-A receptor). Each resulting MoA term defined a specific gene set as the union of its target genes. Since many MoA gene sets are small, sometimes containing only one gene, and often share biology, raw MoA terms were grouped via manual curation into parent sets (*e.g.*, individual integrin heterodimers under a common integrin group), with each parent set defined as the union of its members’ genes. Manual curation was assisted by regular-expression matching of key terms in MoA names to identify candidate groupings, which were then reviewed by hand. Wherever possible, grouping criteria were applied in the following order: (a) clear ligand-receptor signaling pairs were grouped together, and separated into ligand and receptor sets only where each contained ≥ 5 genes; (b) terms outside such a signaling axis (*e.g.*, transporters, ion channels, enzymes, kinases, *etc*.) were grouped by protein family or structural class; (c) terms with neither an axis nor informative family were grouped by pharmacological role or retained as singletons. This produced a two-tiered MoA gene set catalogue: 204 parent MoA sets (median gene count = 5; mean gene count = 8.91; SD = 12.9), of which 61 contained ≥ 2 testable child gene sets directly extracted from ChEMBL v36, yielding 183 child sets (median gene count = 2; mean gene count = 4.82; SD = 6.88).

### Competitive gene set analyses

Competitive gene set analysis was performed with MAGMA v1.10 across all phenotypes with drug and MoA gene sets, using the 1000 Genomes European reference panel. For the MoA sets, all parent (*n* = 204) and child sets (*n* = 183) were run, regardless of whether the parent set was significant. The drug-level and MoA gene set analyses were treated as different testing families. False discovery rate (FDR) correction was applied within each family across all phenotypes, with statistical significance defined at an FDR threshold of *P_FDR_* < 0.05. We correct for the number of unique gene sets tested, since some compounds have identical gene sets.

#### Conditional analyses

Competitive gene set analyses were performed while conditioning on a control gene set containing every gene in the respective set family (drug gene sets, *n* = 5,287; MoA gene sets, *n* = 1,879). This prevented associations solely due to general properties of drug target genes within a given set family. For both the MAGMA *z* and PoPS-based analyses, two competitive gene set analyses were performed: (i) competitive gene set analysis only conditioning on the control gene set (no conditioning on gene expression), and (ii) competitive gene set analysis conditioning on both the control gene set and the average gene expression across (a) all tissues, and (b) all brain tissues. Gene expression values were taken from GTEx v10, and we used the median transcripts per million (TPM) value per gene.

#### MAGMA z-score vs. PoPS-based competitive gene set analysis

Two implementations of competitive gene set analysis were performed in MAGMA: (i) using the standard gene-level *z*-scores computed internally by MAGMA; and (ii) replacing the gene-level *z*-scores for normalized PoPS scores. PoPS scores were computed using gene expression features only, as previous evidence indicates better annotated genes have higher PoPS scores when using the pathway and PPI features^62^.

### Calibration of PoPS-based competitive gene-set test in MAGMA

#### False positive rate of PoPS-based competitive test

MAGMA’s competitive test assumes a gene-level statistic with known null behavior, gene properties and LD structure. These assumptions are not guaranteed to hold for PoPS. To assess calibration, the false positive rate of the PoPS-based competitive gene set test in MAGMA was estimated using 10,000 size-matched null gene sets for both the drug and MoA gene set families. Null gene sets were sampled from all genes in the analysis, as this matched the hypothesis tested by MAGMA’s competitive test, in which a set’s genes are compared against all remaining genes. For each set family, every observed set size was retained once and the remaining sizes were drawn with replacement from a vector of the observed sizes, so that the 10,000 null sets reproduced the empirical size distribution of that family. Genes were then sampled without replacement from the gene pool. The false positive rate was computed as the proportion of null gene sets with a *P*-value < 0.05. We observed for the PoPS-based test that the native *P*-value output by MAGMA was mildly less conservative, with a mean false positive rate of 7.43% compared to the expected 5% (**Supplementary Fig. 1a, b**).

#### PoPS-based competitive test empirical p-value

An empirical *P*-value was adopted for the PoPS-based competitive gene set test to achieve a well calibrated false positive rate. For each phenotype, a null distribution was generated by creating 10.1 million size-matched null gene sets as described above. For each observed gene set, the competitive test statistic (β/SE) was compared against test statistics from the null distribution, and the empirical *P*-value was calculated as (*M*+1)/(*N*+1), where *M* is the number of null sets greater than or equal to the observed statistic. This restored the expected false positive rate (**Supplementary Fig. 1c, d**) and was confirmed with QQ plots per phenotype (**Supplementary Fig. 2**). All PoPS-based results reported above use this empirical *P*-value, while MAGMA *z*-based results use the native MAGMA *P*-value throughout.

#### Screening for systematically biased gene sets

To identify gene sets that were more significant than expected irrespective of phenotype, we ran 1,000 permutations for each phenotype and set family in which PoPS scores were shuffled across genes before analysis. In each permutation, every gene set was assigned the percentile rank of its competitive test statistic relative to all other sets in the same family, which has an expected value of 0.5 in the absence of bias. For each gene set, a two-tailed t-test compared its mean percentile rank across the 1,000 permutations to the expected value of 0.5. *P*-values were corrected using FDR, and all sets reaching significance were excluded from subsequent analyses. Mean percentiles ranged from 0.46 to 0.55 for drug sets and from 0.47 to 0.55 for MoA sets (**Supplementary Fig. 3, 4**). Overall, 15 of 8,151 drug sets (leaving 8,136) and 3 of 387 MoA sets (leaving 384) were removed (Supplementary Table 2) and were excluded.

### Benchmarking recovery of approved drugs and MoAs

To benchmark how MAGMA *z*-score and PoPS-based drug and MoA gene set analyses recover known compounds and biological targets, we performed permutation tests comparing the mean significance rank (drugs ranked by gene-set association *P*-value, most significant first) of drugs and MoAs that are approved for that indication against a null distribution of 10,000 random draws. For drug gene sets, the mean significance rank across approved sets was computed for each phenotype. The null distribution was built by randomly sampling *k* sets and computing the mean significance rank across those sets 10,000 times, where *k* is the true number of approved sets for a given trait. The permutation *P*-value is the proportion of draws with a mean rank at least as low as the observed value.

For MoA gene sets, we performed a block permutation test to account for the parent-child structure of the gene sets, since approval status was annotated to a parent set whenever any of its child sets were approved for the phenotype. The observed statistic was computed by first averaging the significance ranks of the approved sets belonging to the same family and then taking the mean across families. A mechanism of action therefore contributes once regardless of how many of its sets are approved, preventing families where both the parent and one or more children are approved from being counted multiple times. Sets from those families that were not approved for the phenotype remained in the ranking and in the null pool but did not contribute to the family average. The null was constructed by drawing sets with the same structure as the observed approved sets: if the approved sets for a phenotype comprised one parent-child pair and two single sets, then every random draw consisted of two members of a randomly chosen family and two randomly drawn single sets. The permutation *P*-value is the proportion of the 10,000 draws with a mean rank at least as low as the observed value.

To define what constitutes “approval” we used the following EFO indication labels with a clinical maximum phase of 4.0 (indicating approval) from ChEMBL: ADHD = “attention deficit hyperactivity disorder”; AD = “Alzheimer disease”; asthma = “asthma”; BIP = “bipolar disorder”, “bipolar I disorder”, “bipolar II disorder”; CAD = “coronary artery disease”; IBD = “Crohn’s disease”, “ulcerative colitis”; LDL = “Hypercholesterolemia”, “familial hypercholesterolemia”, “homozygous familial hypercholesterolemia”; MDD = “major depressive disorder”; SCZ = “schizophrenia”, “schizoaffective disorder”; T2D = “type 2 diabetes mellitus”. Significance was defined as FDR-corrected permuted *P* < 0.05 for MAGMA *z* and PoPS-based analyses separately.

### Benchmarking attained clinical development stage

We assessed if higher gene set significance was associated with a compound set having a higher clinical development phase. For each phenotype we extracted compounds (*n* = 1,360) that mapped to the EFO indication labels for our phenotypes (see previous) and had a clinical phase annotation of 1.0 to 4.0. Compounds were grouped by gene set content (i.e., drugs with identical gene targets were treated as a single set), resulting in 1,266 unique sets. The maximum development phase among drugs with identical sets was used as that set’s clinical phase. The association between gene set significance and attained clinical stage was estimated using Spearman’s rho between the -log_10_(*P*) and attained clinical phase (1, 2, 3, or 4) for each phenotype, and *P*-values were FDR-corrected. Spearman rho statistics were transformed to Fisher’s *z* using the inverse hyperbolic tangent, and 95% confidence intervals were constructed using *Z* ± 1.96 × sqrt(1.06 / (*n* – 3)), using the variance form appropriate for Spearman’s rho, and back transformed by the hyperbolic tangent. Phenotype-specific estimates were combined using a random-effects meta-analysis on the Fisher *z* scale, with between-study variance estimated by the Paule-Mandel method and inference by the Hartung-Knapp adjustment (safeguarded so that the standard error never fell below the ordinary random-effects value). Table 2 displays the pooled rho with its 95% confidence interval, 95% prediction interval, tau^2^, I^2^, and Cochran’s Q. All phenotypes had ≥ 10 effective gene sets spanning ≥ 2 clinical stages. For display only, gene set significance was re-scaled to within-phenotype percentile, which is rank preserving and alters no reported statistic.

### Drug class enrichment analysis

To test whether drugs of the same therapeutic class showed stronger drug gene-set associations than other drugs, compounds were grouped by their ATC level 3 code (ChEMBL v36). Every class containing at least five drugs with a testable gene set was examined for enrichment of genetic signals using a multiple linear regression model to estimate the effect of drug group membership on MAGMA t-statistics. Given that the MAGMA t-statistic is a measure of the degree to which a drug gene set is enriched for genetic signal for a phenotype, this allows us to test whether drugs in that drug group exhibited greater enrichment than other drugs.

For each phenotype and implementation (MAGMA *z* or PoPS-based), let us denote the vector of drug gene-set t-statistics as *Y* and encoding a binary indicator variable *G*_j_ for drug group *j*, scored 1 for drugs in that drug group and 0 otherwise, we fit the model *Y* = *β*_0_ + *Cβ_C_* + *G*_j_*β_G_* + *ε* = *Xβ* + *ε*. Here, *C* is a covariate matrix containing the drug gene-set size as well as the log of the drug gene-set size, and *ε* is the residual term with distribution *ε* ∼ *MVN*(0, *σ*^2^*S*), where *S* is the sampling correlation matrix of the MAGMA t-statistics. Combining the intercept and predictor variables in *X*, the model can be estimated using Generalized Least Squares, yielding the estimate *β̂* _=_ (*_X_^T^_S_*^−1^*_X_*)^−1^*_X_^T^_S_*^−1^*_Y_*, with sampling covariance matrix *W* = *σ̂*^2^(*X^T^S*^−1^*X*)^−1^. For the residual variance we have the estimate 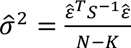 with *ɛ̂* = *Y* − *Xβ̂* and with *N* the number of drugs and *K* = 4 the number of *N*−*K* parameters in the regression equation. For each drug group *j*, we test the null hypothesis *H* : *β* = 0 against the one-sided alternative *H* : *β* > 0 using the test statistic 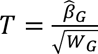 where *W_G_* is the diagonal element of *W* corresponding to *β̂G* and *T* has a t-distribution with *N* − *K* degrees of freedom.

Each model was fitted twice: with *S*, our primary test, and with *S* replaced by the identity matrix (ordinary least squares), which treats drugs as independent. Because drugs that share targets have correlated test statistics regardless of how they act on those targets, the second model is expected to be less conservative for classes whose members share targets; we report it to show which class-level signals depend on target sharing. This gave four analyses per phenotype: MAGMA *z*-score and PoPS-based statistics, each with and without *S*. Within each analysis, *P*-values were FDR corrected across all ten phenotypes and all classes tested (1,355 tests per analysis), and classes with *P_FDR_* < 0.05 were considered significant.

### Concordance between implementations and gene set families

Agreement between the MAGMA *z* and PoPS-based implementations was estimated per phenotype as the Pearson correlation of the competitive test statistics (β/SE), computed separately for drug and MoA sets.

To compare the two gene set families, each drug gene set with a ChEMBL MoA annotation was paired with its annotated mechanism. If a drug carried both a parent and child annotation, the child mechanism was used. Drugs without MoA annotations were excluded from the pairing but retained when calculating the proportion of significant drug sets lacking a corresponding mechanism. For each phenotype and implementation, convergence was assessed in two ways. First, study-wide significant sets were classified as convergent (a significant MoA set with at least one significant annotated drug set, or vice versa), unique to one family (or, for drug sets, lacking a MoA annotation altogether), and the proportion of each category was calculated. Second, the competitive test statistic of each MoA gene set was correlated with the mean test statistic of its annotated compounds using a Peason correlation.

## Data availability

The drug and mechanism-of-action gene sets created for this study are available through the DRUGSETS tool (https://github.com/nybell/drugsets). GWAS summary statistics used for this study are available as follows: ADHD, BIP, MDD, and SCZ, pgc.unc.edu/for-researchers/download-results/; asthma, ebi.ac.uk/gwas/publications/32296059; LDL, csg.sph.umich.edu/willer/public/glgc-lipids2021/; CAD, kp4cd.org/node/348; IBD, bio-computing.hrbmu.edu.cn/posgwaser/; T2D, diagram-consortium.org/. AD summary statistics were obtained directly from the study authors^37^.

## Code availability

DRUGSETS is available as an open-source Python package located at https://github.com/nybell/drugsets and is being added to the FUMA platform (https://fuma.ctglab.nl/). Analysis and figure-generation scripts are available at Zenodo: https://doi.org/10.5281/zenodo.23587828.

## Acknowledgements

This project was supported by the European Union’s Horizon 2020 research and innovation programme under grant agreement No 964874 (REALMENT), and by the Netherlands Organization for Scientific Research - Gravitation project ‘BRAINSCAPES: A Roadmap from Neurogenetics to Neurobiology’ (024.004.012). DP and EU were funded by the European Research Council advanced grant ‘From GWAS to Function’ (ERC-2018-ADG 834057). EvW is funded via the Ministry of Education, Culture, and Science in The Netherlands. NYB, EU, MS, EvW, and CdL have no conflicts of interest to declare. DP is a consultant for Temple Private Ltd. KH was supported by a Humboldt Research Fellowship from the Alexander von Humboldt Foundation and is now an employee of Bayer AG.

