## Supplemental Figures for "Drug Gene-Set Analysis Identifies Established and Emerging Therapeutic Mechanisms"

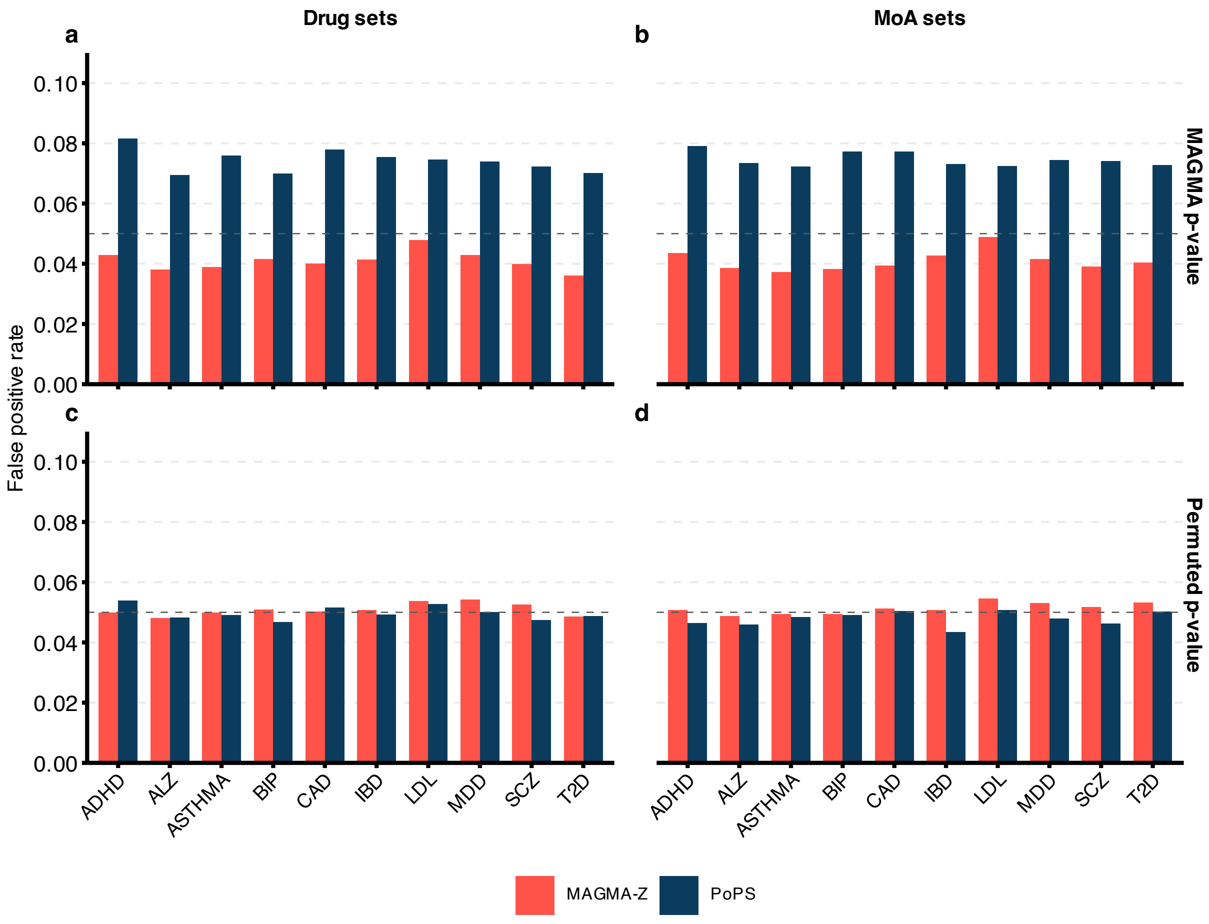


**Supplemental Fig. 1 | False positive rates of MAGMA-z and PoPS-based gene set analyses. (a-d)** Bar plots depicting the false positive rates of null competitive gene set analyses conditioned on gene expression (see Methods). Results are shown for the native MAGMA *P*-value (top row; **a**, **b**) and permutation-based empirical *P* (bottom row; **c**, **d**; see Methods for details). The left column (**a**, **c**) displays results for drug gene sets, while the right column (**b**, **d**) shows results for MoA sets. Color denotes method used: vermilion = MAGMA; indigo = PoPS.

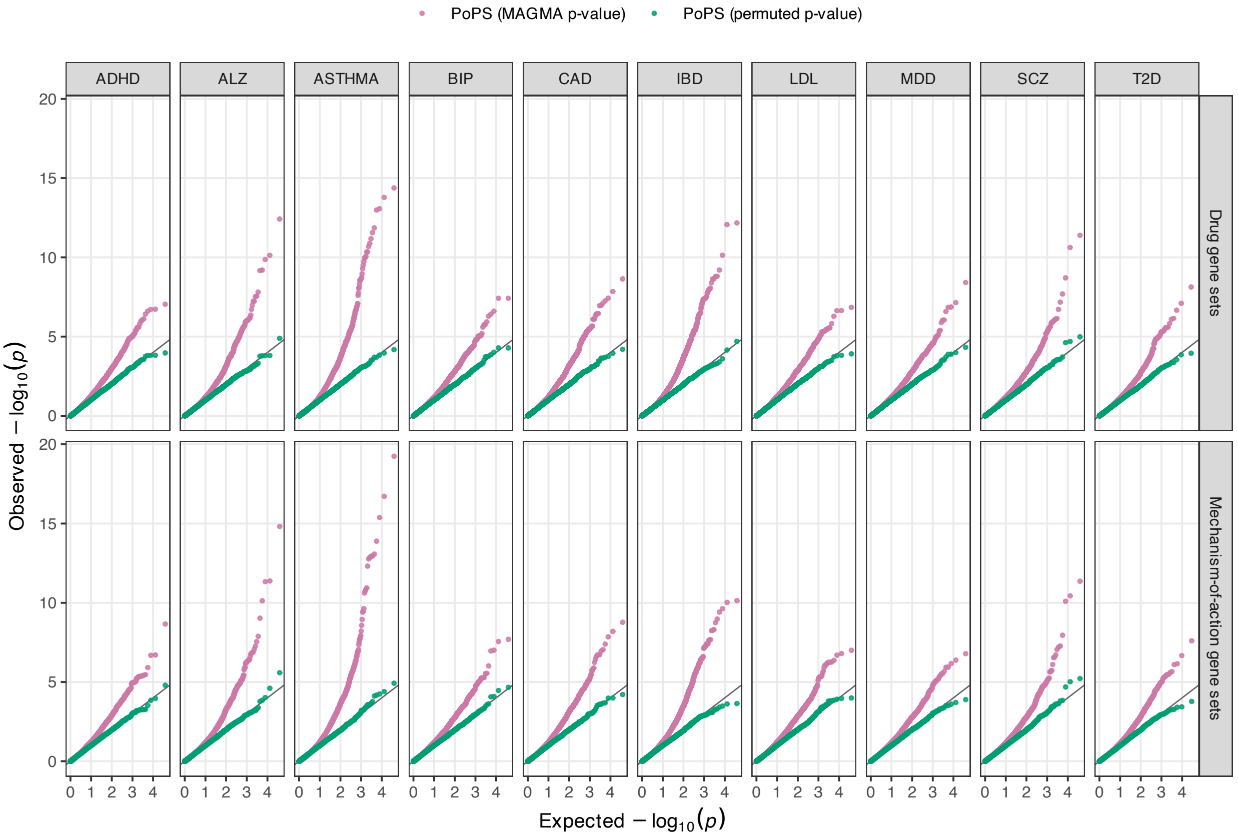


**Supplemental Fig. 2 | QQ plots of null gene-set *P*-values across ten phenotypes for PoPS-based gene set analysis.** Quantile-quantile plots of *P*-values from 10 million size-matched null gene sets, analysed with the PoPS-based implementation of MAGMA's competitive gene-set test. Columns show the ten phenotypes, with the upper row showing drug gene set results, and the bottom row showing the MoA gene set results. Each panel plots the observed against expected -$\log_{10}(P)$ for MAGMA's native *P* (pink) and the empirical permutation-based *P* (green). The diagonal marks the expectation under the null.

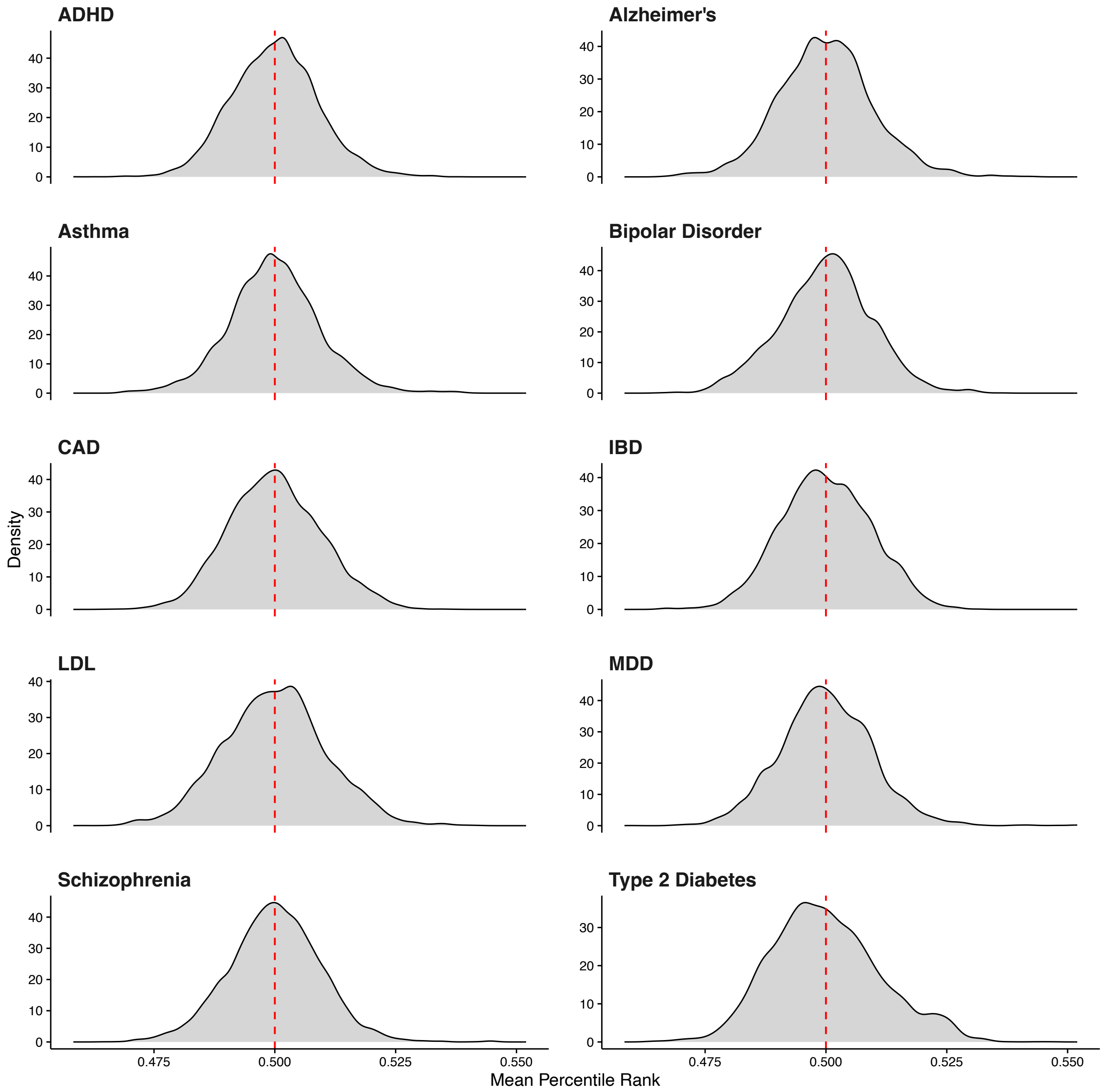


**Supplemental Fig. 3 | Distributions of mean percentile ranks for drug gene sets.** Distributions of the mean percentile rank of the competitive test statistic (β/SE) for each drug gene set, computed across 1,000 permutations. For each permutation, PoPS scores were randomly shuffled prior to analysis. Panels show the ten phenotypes, with a range across all from 0.46 to 0.55. The red dashed line marks 0.5, the expected value for each. Sets departing significantly from 0.5 by two-sided t-test after FDR correction were excluded from subsequent analyses (15 of *n* = 8,151 drug gene sets; **Supplementary Table 2**).

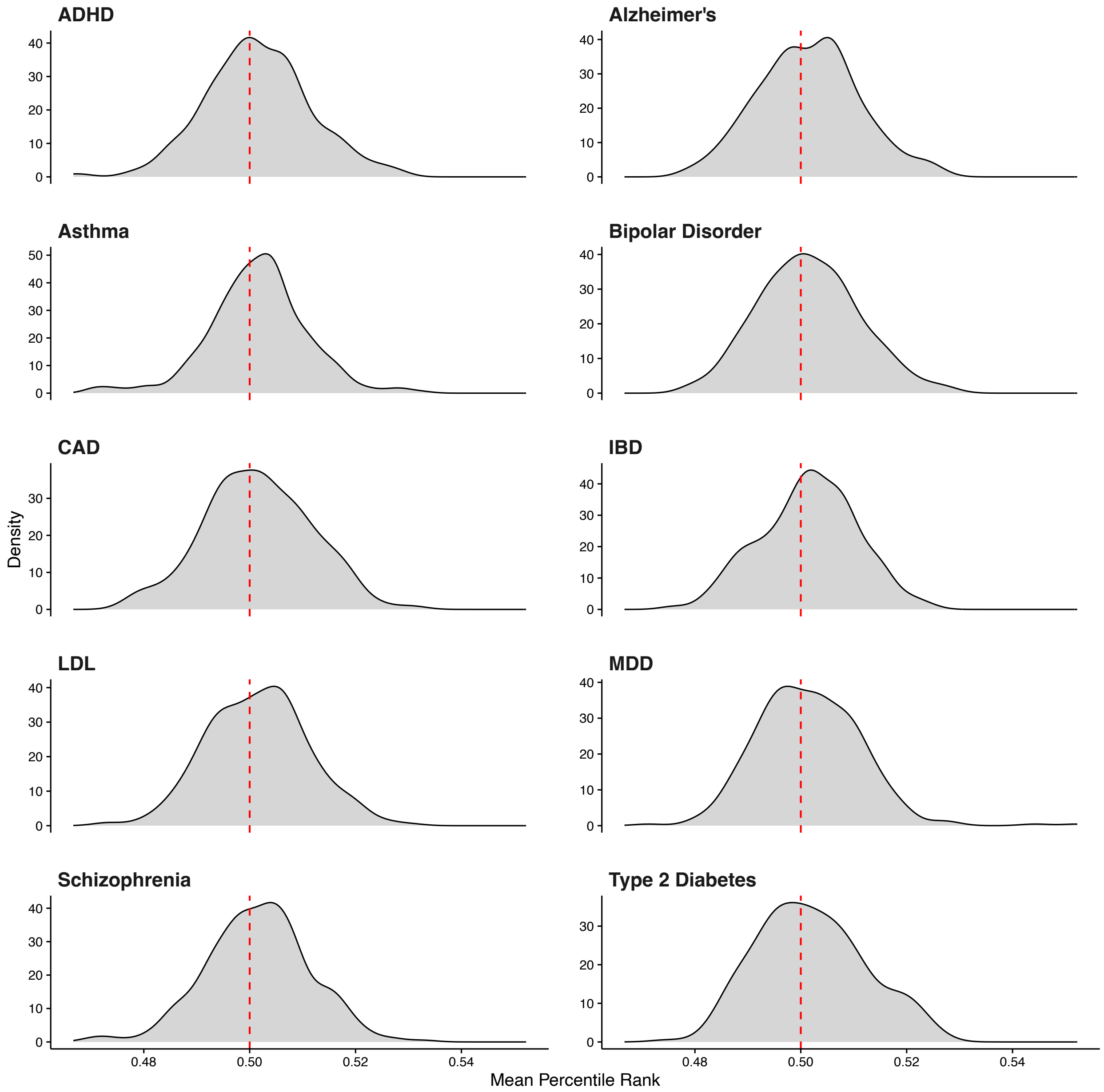


**Supplemental Fig. 4 | Distributions of mean percentile ranks for MoA gene sets.** Distributions of the mean percentile rank of the competitive test statistic (β/SE) for each MoA gene set, comparable to **Supplementary Fig. 2**. For MoA sets, mean percentile ranks ranged from 0.47 to 0.55, and sets departing significantly from 0.5 by two-sided t-test after FDR correction were excluded from subsequent analyses (3 of *n* = 387 MoA gene sets; **Supplementary Table 2**).

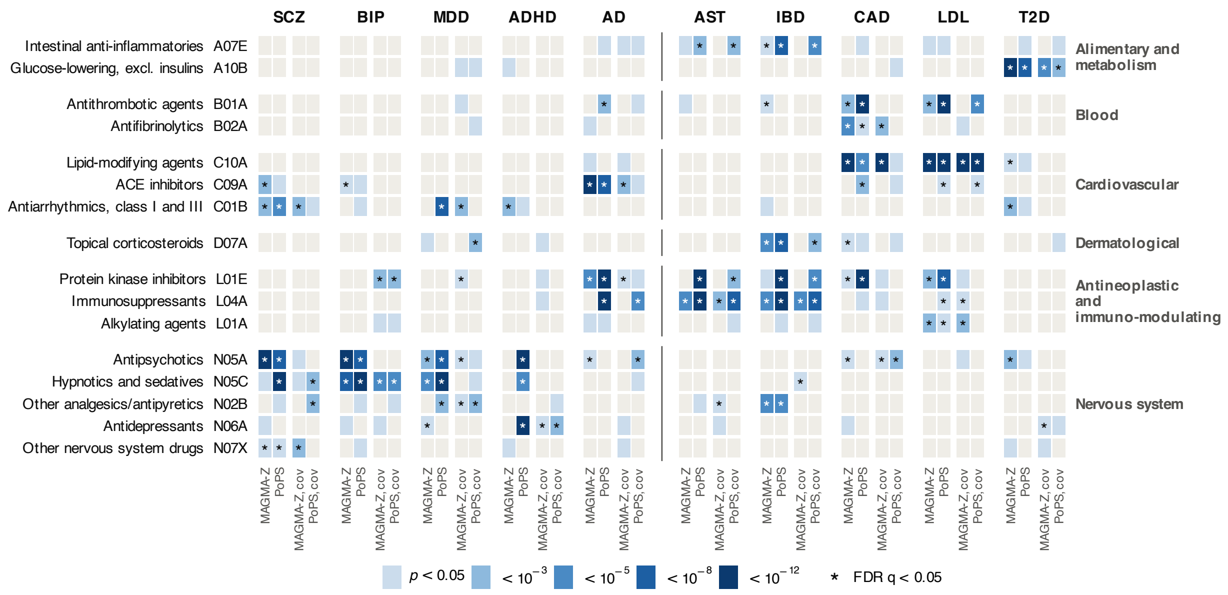


**Supplementary Figure 5 | ATC level 3 class enrichment results.** Heatmap displaying enrichment of phenotype-ATC level 3 class pairs that are significant in at least three of the four analysis types. The X axis labels (top) show phenotype names and analysis types (bottom), and ATC3 codes are on the Y axis (left), with the ATC level 1 label on the right. * = study-wide $P_{FDR} < 0.05$.
