## Supplemental Note 1 for "Drug Gene-Set Analysis Identifies Established and Emerging Therapeutic Mechanisms"

### Supplementary Note 1: Drug class enrichment analyses

Drug class enrichment analysis offers an alternative to the mechanism-of-action (MoA) gene set analysis for testing drug groups. While the MoA gene set analysis can prioritize multiple drugs by identifying a biological target, the drug class enrichment tests whether groups of compounds in the same ATC level 3 class (ATC3) have, on average, stronger drug gene-set associations compared to drugs not in the group. This note explains how to read the drug class enrichment analysis (Supplementary Fig. 4; Supplemental Table 6). The model and multiple-testing correction are described in Methods.

Each ATC3 class was tested with and without modelling the correlation between drugs. Drugs whose gene sets overlap will have correlated test statistics, since both are computed from the same gene-level statistics. This correlation exists whether or not the drugs act on the shared gene targets in the same way, since the gene-set test uses gene membership only. Therefore, the analysis that models the correlation between drug gene-set t-statistics from MAGMA is used as the primary. However, we also test drug group enrichment without modelling the correlation between drug gene sets, and we make both options available in the DRUGSETS tool. The uncorrected model treats drugs as independent and is therefore less conservative when drug class members share gene targets; we report it to show which signals depend on that sharing.

We observe 150 phenotype-ATC3 class pairs that were enriched in at least one of the MAGMA *z*-based results after study-wide FDR correction: 20 were significant both with and without correction for correlated drug gene-set t-statistics, 79 were significant only without correction, and 51 were significant only with correction. For the PoPS-based results, of the 146 phenotype-ATC3 class pairs that were significant in at least one analysis, 21 were significant both with and without correction, 98 were significant only without the correction, and 27 only with the correction. Five phenotype-class pairs were significantly enriched across all analysis types: lipid modifying agents (C10A) for LDL cholesterol, blood glucose-lowering drugs (A10B) for T2D, immunosuppressants (L04A) for IBD and asthma, and hypnotics and sedatives (N05C) for BIP (Supplemental Fig. 4;

Supplementary Table 6). Three of these are established treatment classes for the phenotype tested (C10A for LDL cholesterol, A10B for T2D and L04A for IBD), with immunosuppressants for asthma and hypnotics and sedatives for BIP being the exceptions. Fourteen phenotype-ATC3 class pairs showed study-wide enrichment for three of the four analysis types, including lipid modifying agents (C10A) for CAD, intestinal anti-inflammatory agents (A07E) for IBD, ACE inhibitors (C09A) for AD, and anti-depressants (N06A) for ADHD. Lastly, two classes were significant for seven of the ten phenotypes in at least one analysis: protein kinase inhibitors (L01E) and anti-psychotics (N05A). This possibly indicates that these drug groups are enriched for gene targets with pleiotropic rather than disease-specific effects.

Overall, due to the shared inputs between the two covariance analysis options, and the correlated nature of the inputs between MAGMA *z*-score and PoPS-based results, concordance between the results indicates reliability of the results rather than independent replication. ATC3 classes that are significant without the covariance correction, but that are insignificant with, may reflect a small number of strongly associated genes shared across drugs in the class.
